# Clinical characteristics of 2019 novel coronavirus infection in China

**DOI:** 10.1101/2020.02.06.20020974

**Authors:** Wei-jie Guan, Zheng-yi Ni, Yu Hu, Wen-hua Liang, Chun-quan Ou, Jian-xing He, Lei Liu, Hong Shan, Chun-liang Lei, David S.C. Hui, Bin Du, Lan-juan Li, Guang Zeng, Kwok-Yung Yuen, Ru-chong Chen, Chun-li Tang, Tao Wang, Ping-yan Chen, Jie Xiang, Shi-yue Li, Jin-lin Wang, Zi-jing Liang, Yi-xiang Peng, Li Wei, Yong Liu, Ya-hua Hu, Peng Peng, Jian-ming Wang, Ji-yang Liu, Zhong Chen, Gang Li, Zhi-jian Zheng, Shao-qin Qiu, Jie Luo, Chang-jiang Ye, Shao-yong Zhu, Nan-shan Zhong, on behalf of China Medical Treatment Expert Group for 2019-nCoV

## Abstract

**Background:** Since December 2019, acute respiratory disease (ARD) due to 2019 novel coronavirus (2019-nCoV) emerged in Wuhan city and rapidly spread throughout China. We sought to delineate the clinical characteristics of these cases.

**Methods:** We extracted the data on 1,099 patients with laboratory-confirmed 2019-nCoV ARD from 552 hospitals in 31 provinces/provincial municipalities through January 29^th^, 2020.

**Results:** The median age was 47.0 years, and 41.90% were females. Only 1.18% of patients had a direct contact with wildlife, whereas 31.30% had been to Wuhan and 71.80% had contacted with people from Wuhan. Fever (87.9%) and cough (67.7%) were the most common symptoms. Diarrhea is uncommon. The median incubation period was 3.0 days (range, 0 to 24.0 days). On admission, ground-glass opacity was the typical radiological finding on chest computed tomography (50.00%). Significantly more severe cases were diagnosed by symptoms plus reverse-transcriptase polymerase-chain-reaction without abnormal radiological findings than non-severe cases (23.87% vs. 5.20%, *P*<0.001). Lymphopenia was observed in 82.1% of patients. 55 patients (5.00%) were admitted to intensive care unit and 15 (1.36%) succumbed. Severe pneumonia was independently associated with either the admission to intensive care unit, mechanical ventilation, or death in multivariate competing-risk model (sub-distribution hazards ratio, 9.80; 95% confidence interval, 4.06 to 23.67).

**Conclusions:** The 2019-nCoV epidemic spreads rapidly by human-to-human transmission. Normal radiologic findings are present among some patients with 2019-nCoV infection. The disease severity (including oxygen saturation, respiratory rate, blood leukocyte/lymphocyte count and chest X-ray/CT manifestations) predict poor clinical outcomes.

## Introduction

In early December 2019, the first pneumonia cases of unknown origins were identified in Wuhan city, Hubei province, China [1]. High-throughput sequencing has revealed a novel betacoronavirus that is currently named 2019 novel coronavirus (2019-nCoV) [2], which resembled severe acute respiratory syndrome coronavirus (SARS-CoV) [3]. The 2019-nCoV is the seventh member of enveloped RNA coronavirus (subgenus *sarbecovirus, Orthocoronavirinae* subfamily) [3]. Evidence pointing to the person-to-person transmission in hospital and family settings has been accumulating [4-8].

The World Health Organization has recently declared the 2019-nCoV a public health emergency of international concern [9]. As of February 5^th^, 2020, 24,554 laboratory-confirmed cases have been documented globally (i.e., the USA, Vietnam, Germany) [5,6,9,10]. 28,018 laboratory-confirmed cases and 563 death cases in China as of February 6^th^, 2020 [11]. Despite the rapid spread worldwide, the clinical characteristics of 2019-nCoV acute respiratory disease (ARD) remain largely unclear. In two recent studies documenting the clinical manifestations of 41 and 99 patients respectively with laboratory-confirmed 2019-nCoV ARD who were admitted to Wuhan, the severity of some cases with 2019-nCoV ARD mimicked that of SARS-CoV [1,12]. Given the rapid spread of 2019-nCoV, an updated analysis with significantly larger sample sizes by incorporating cases throughout China is urgently warranted. This will not only identify the defining epidemiological and clinical characteristics with greater precision, but also unravel the risk factors associated with mortality. Here, by collecting the data from 1,099 laboratory-confirmed cases, we sought to provide an up-to-date delineation of the epidemiological and clinical characteristics of patients with 2019-nCoV ARD throughout mainland China.

## Methods

### Data sources

We performed a retrospective study on the clinical characteristics of laboratory-confirmed cases with 2019-nCoV ARD. The initial cases were diagnosed as having ‘pneumonia of unknown etiology’, based on the clinical manifestations and chest radiology after exclusion of the common bacteria or viruses associated with community-acquired pneumonia. Suspected cases were identified as having fever or respiratory symptoms, and a history of exposure to wildlife in Wuhan seafood market, a travel history or contact with people from Wuhan within 2 weeks [13]. Cases were diagnosed based on the WHO interim guidance [14]. A confirmed case with 2019-nCoV ARD was defined as a positive result to high-throughput sequencing or real-time reverse-transcriptase polymerase-chain-reaction (RT-PCR) assay for nasal and pharyngeal swab specimens [1]. Only the laboratory-confirmed cases were included the analysis. The incubation period was defined as the duration from the contact of the transmission source to the onset of symptoms. The study was approved by the National Health Commission and the institutional board of each participating site. Written informed consent was waived in light of the urgent need to collect clinical data.

The epidemiological characteristics (including recent exposure history), clinical symptoms and signs and laboratory findings were extracted from electronic medical records. Radiologic assessments included chest X-ray or computed tomography. Laboratory assessments consisted of complete blood count, blood chemistry, coagulation test, liver and renal function, electrolytes, C-reactive protein, procalcitonin, lactate dehydrogenase and creatine kinase. The severity of 2019-nCoV ARD was defined based on the international guidelines for community-acquired pneumonia [15].

The primary composite endpoint was the admission to intensive care unit (ICU), or mechanical ventilation, or death. Secondary endpoints comprised mortality rate, the time from symptom onset to the composite endpoint and each of its component. Because clinical observations were still ongoing, fixed time frame (i.e. within 28 days) was not applied to these endpoints.

All medical records were copied and sent to the data processing center in Guangzhou, under the coordination of the National Health Commission. A team of experienced respiratory clinicians reviewed and abstracted the data. Data were entered into a computerized database and cross-checked. If the core data were missing, requests of clarification were immediately sent to the coordinators who subsequently contacted the attending clinicians. The definition of exposure to wildlife, acute respiratory distress syndrome (ARDS), pneumonia, acute kidney failure, acute heart failure and rhabdomyolysis are provided in the *Supplementary Appendix*.

### Laboratory confirmation

Laboratory confirmation of the 2019-nCoV was achieved through the concerted efforts of the Chinese Center for Disease Prevention and Control (CDC), the Chinese Academy of Medical Science, Academy of Military Medical Sciences, and Wuhan Institute of Virology. The RT-PCR assay was conducted in accordance with the protocol established by the World Health Organization [16]. Further details are available in the *Supplementary Appendix*.

### Statistical analysis

Continuous variables were expressed as the means and standard deviations or medians and interquartile ranges (IQR) as appropriate. Categorical variables were summarized as the counts and percentages in each category. We grouped patients into severe and non-severe 2019-nCoV ARD according to the American Thoracic Society guideline on admission [15]. Wilcoxon rank-sum tests were applied to continuous variables, chi-square tests and Fisher’s exact tests were used for categorical variables as appropriate. The risk of composite endpoints among hospitalized cases and the potential risk factors were analyzed using Fine-Gray competing-risk models in which recovery is a competing risk. The proportional hazard Cox model was used in sensitivity analyses. The candidate risk factors included an exposure history, greater age, abnormal radiologic and laboratory findings, and the development of complications. We fitted univariate models with a single candidate variable once at a time. The statistically significant risk factors, sex, and smoking status were included into the final models. The sub-distribution hazards ratio (SDHR) along with the 95% confidence interval (95%CI) were reported. All analyses were conducted with R software version 3.6.2 (R Foundation for Statistical Computing). Distribution map was plotted using ArcGis version 10.2.2.

## Results

### Demographic and clinical characteristics

Of all 1,324 patients recruited as of January 29^th^, 222 (16.8%) had a suspected diagnosis and were therefore excluded. The core data sets (including clinical outcomes and symptoms) of 3 patients were lacking due to the incompleteness of original reports, hence this report delineates 1,099 patients with 2019-nCoV ARD from 552 hospitals in 31 provinces/province-level municipalities (**Fig. 1**).

**Figure 1.**
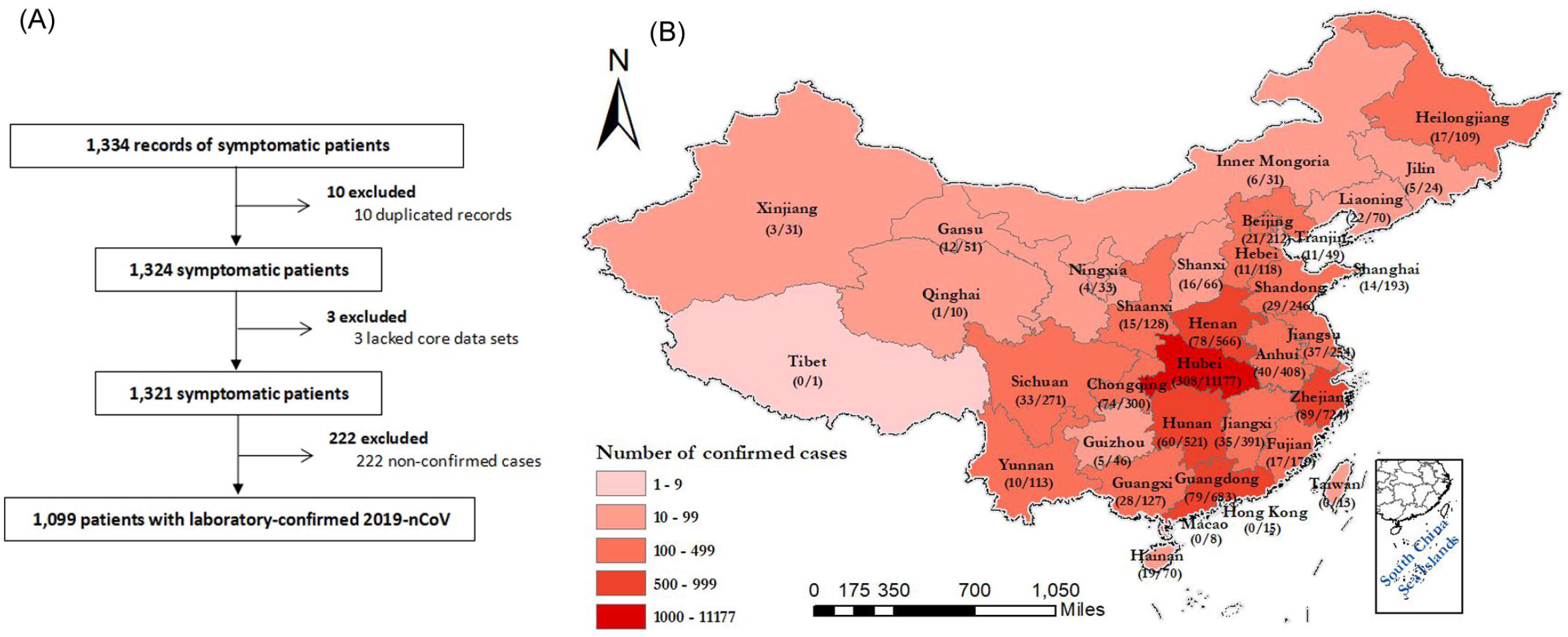
Patient recruitment flowchart and the distribution of patients across China. Figure 1-A. Patient recruitment flowchart Figure 1-B. The distribution of laboratory-confirmed cases throughout China Shown are the official statistics of all documented laboratory-confirmed cases throughout China according to the National Health Commission (as of February 4^th^, 2020).

The demographic and clinical characteristics are shown in **Table 1**. 2.09% were healthcare workers. A history of contact with wildlife, recent travel to Wuhan, and contact with people from Wuhan was documented in 1.18%, 31.30% and 71.80% of patients, respectively. 483 (43.95%) patients were local residents of Wuhan. 26.0% of patients outside of Wuhan did not have a recent travel to Wuhan or contact with people from Wuhan. The median incubation period was 3.0 days (range, 0 to 24.0).

**Table 1.**
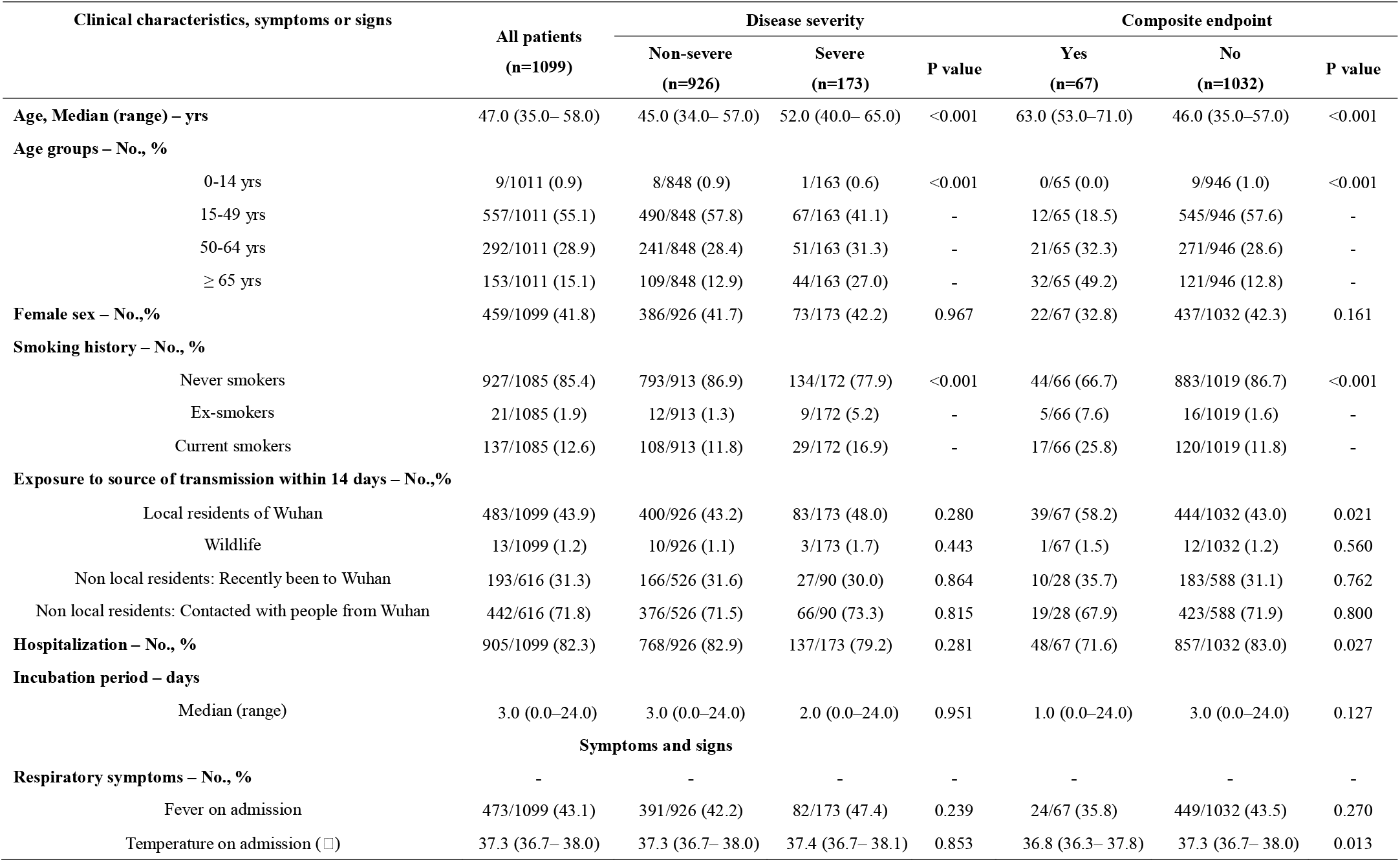
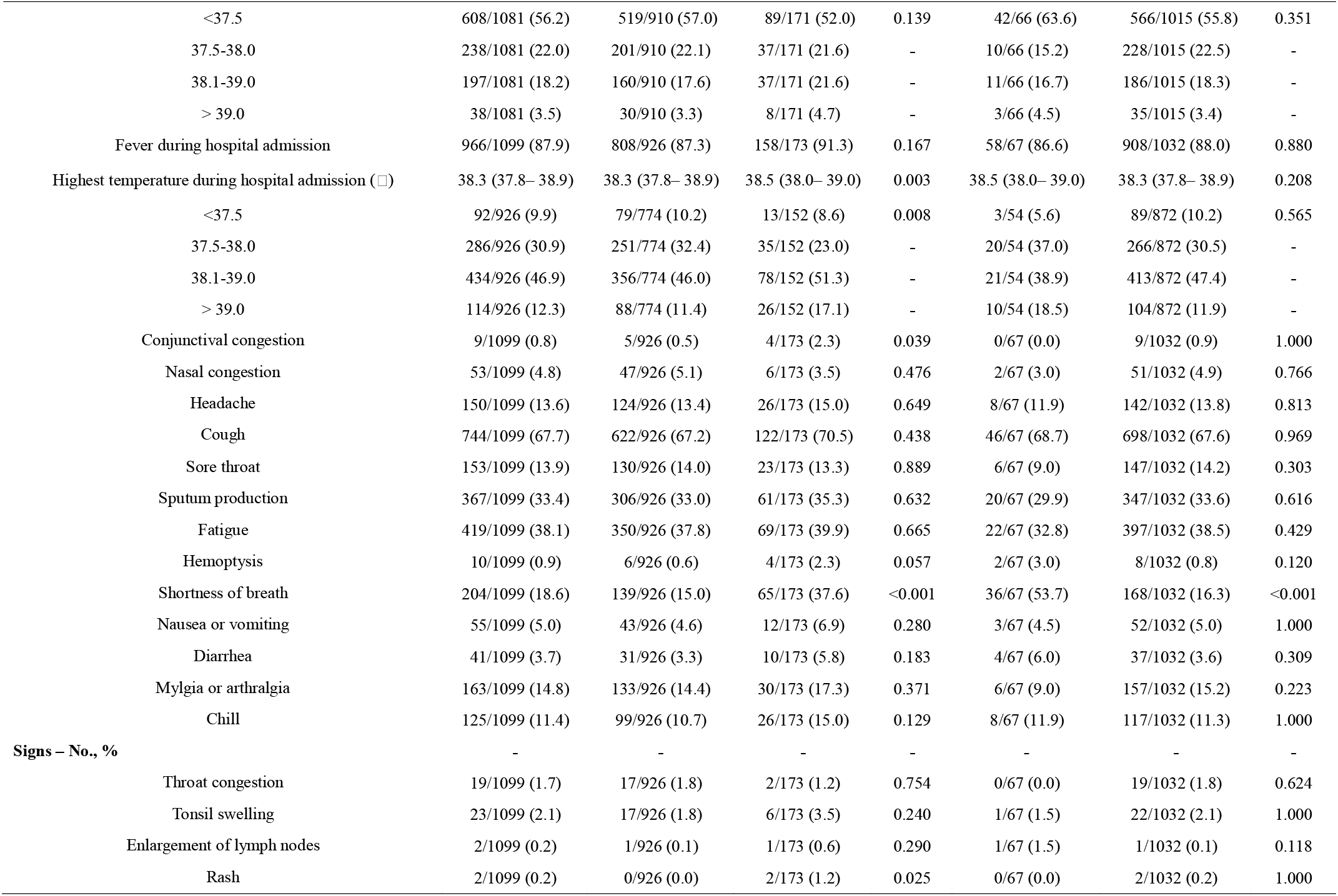
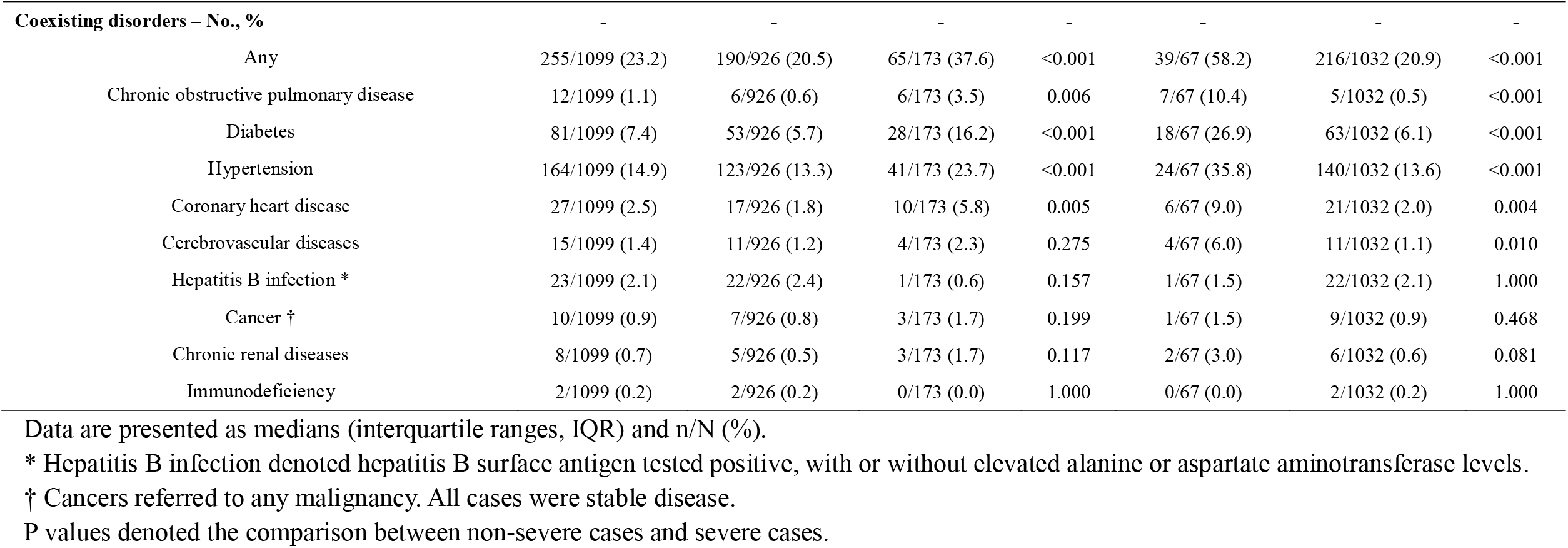
Clinical characteristics of 1,099 patients with 2019-nCoV ARD.

The median age was 47.0 years (IQR, 35.0 to 58.0), and 41.9% were females. 2019-nCoV ARD was diagnosed throughout the whole spectrum of age. 0.9% of patients were aged below 15 years. Fever (87.9%) and cough (67.7%) were the most common symptoms, whereas diarrhea (3.7%) and vomiting (5.0%) were rare. 25.2% of patients had at least one underlying disorder (i.e., hypertension, chronic obstructive pulmonary disease). On admission, 926 and 173 patients were categorized into non-severe and severe subgroups, respectively. The age differed significantly between the two groups (mean difference, 7.0, 95%CI, 4.4 to 9.6). Moreover, any underlying disorder was significantly more common in severe cases as compared with non-severe cases (38.2% vs. 22.5%, *P*<0.001). There were, however, no marked differences in the exposure history between the two groups (all *P*>0.05).

### Radiologic and laboratory findings at presentation

**Table 2** shows the radiologic and laboratory findings on admission. Of 840 patients who underwent chest computed tomography on admission, 76.4% manifested as pneumonia. The most common patterns on chest computed tomography were ground-glass opacity (50.0%) and bilateral patchy shadowing (46.0%). **Figure E1** in the *Supplementary Appendix* demonstrates the representative radiologic findings of two patients with non-severe 2019-nCoV ARD and another two patients with severe 2019-nCoV ARD. Despite these predominant manifestations, 221 out of 926 (23.87%) in severe cases compared with 9 out of 173 non-severe cases (5.20%) who had no abnormal radiological findings were diagnosed by symptoms plus RT-PCR positive findings (*P*<0.001). Severe cases yielded more prominent radiologic abnormalities on chest X-ray and computed tomography than non-severe cases (all *P*<0.05).

**Table 2.**
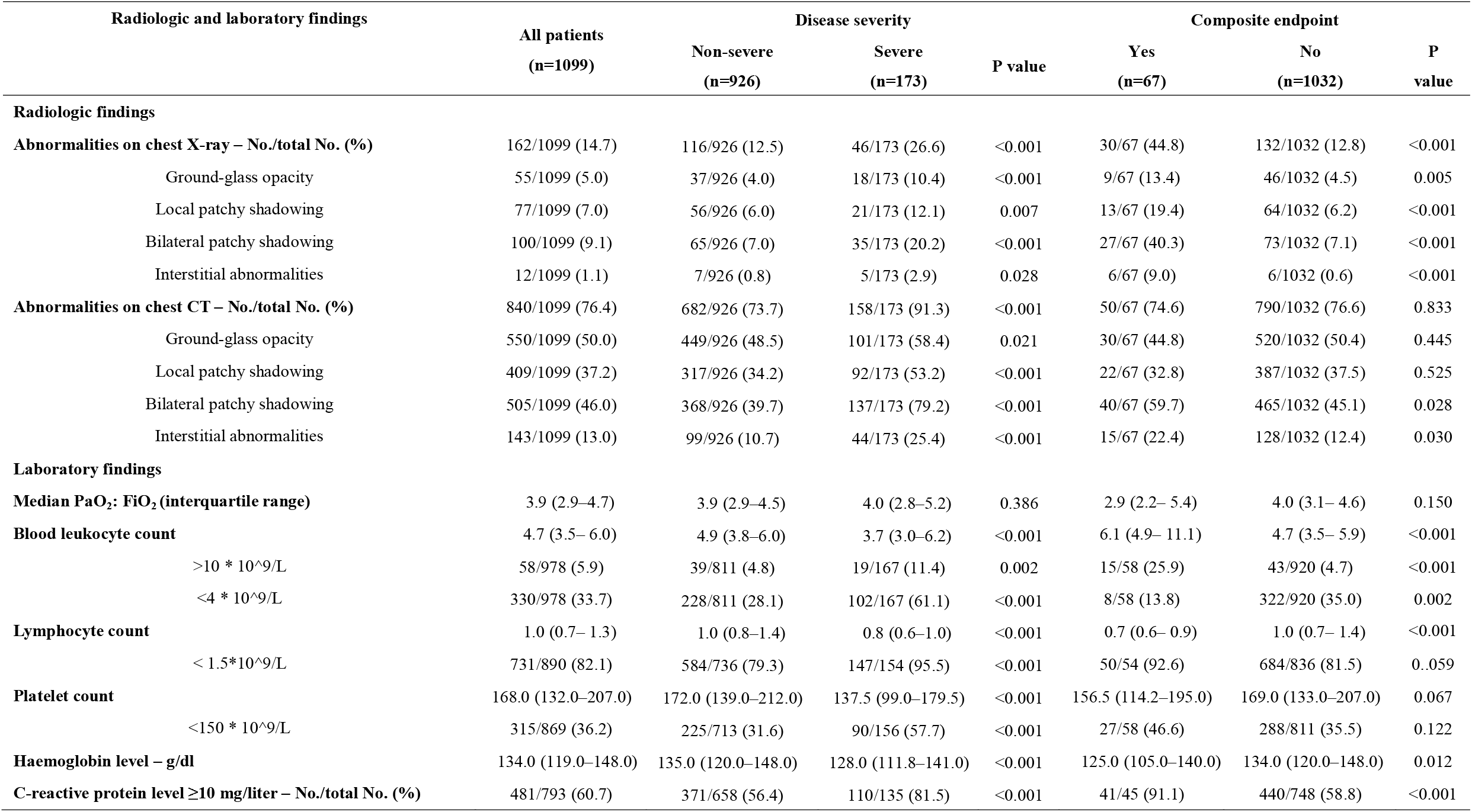
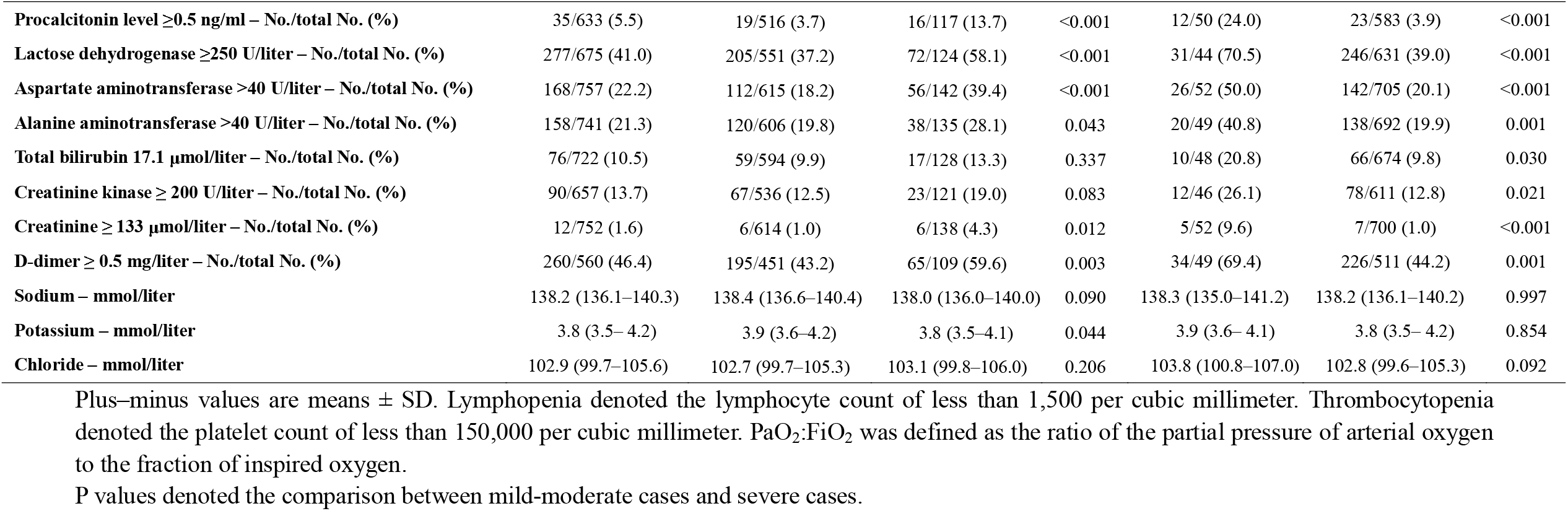
Radiographic and laboratory findings of 1,099 patients with 2019-nCoV ARD.

On admission, 82.1% and 36.2% of patients had lymphopenia and thrombocytopenia, respectively. Overall, leukopenia was observed in 33.7% of patients. Most patients demonstrated elevated levels of C-reactive protein, but elevated levels of alanine aminotransferase, aspartate aminotransferase, creatine kinase and D-dimer were less common. Severe cases had more prominent laboratory abnormalities (i.e., leukopenia, lymphopenia, thrombocytopenia, elevated C-reactive protein levels) as compared with non-severe cases (all *P*<0.05).

### Treatment and complications

Overall, oxygen therapy, mechanical ventilation, intravenous antibiotics and oseltamivir therapy were initiated in 38.0%, 6.1%, 57.5% and 35.8% of patients, respectively. All these therapies were initiated in significantly higher percentages of severe cases (all *P*<0.05). Significantly more severe cases received mechanical ventilation (non-invasive: 32.37% vs. 0%, *P*<0.001; invasive: 13.87% vs. 0%, *P*<0.001) as compared with non-severe cases. Systemic corticosteroid was given to 18.6% of cases and more so in the severe group than the non-severe patients (44.5% vs 13.7%, p<0.001). Moreover, extracorporeal membrane oxygenation was adopted in 5 severe cases but none in non-severe cases (*P*<0.001).

During hospital admission, the most common complication was pneumonia (79.1%), followed by ARDS (3.37%) and shock (1.00%). Severe cases yielded significantly higher rates of any complication as compared with non-severe cases (94.8% vs. 72.2%, *P*<0.001) (**Table 3**).

**Table 3.** Complications, treatment and outcomes of 1,099 patients with 2019-nCoV ARD.

| Characteristics | All patients<br>(n=1099) | Disease severity |  |  | Composite endpoint |  |  |
| --- | --- | --- | --- | --- | --- | --- | --- |
|  |  | Non-severe<br>(n=926) | Severe<br>(n=173) | P value | Yes<br>(n=67) | No<br>(n=1032) | P value |
| Complications – No., % |  |  |  |  |  |  |  |
| Septic shock | 11/1099 (1.0) | 0/926 (0.0) | 11/173 (6.4) | <0.001 | 9/67 (13.4) | 2/1032 (0.2) | <0.001 |
| Acute respiratory distress syndrome | 37/1099 (3.4) | 10/926 (1.1) | 27/173 (15.6) | <0.001 | 27/67 (40.3) | 10/1032 (1.0) | <0.001 |
| Acute kidney injury | 6/1099 (0.5) | 1/926 (0.1) | 5/173 (2.9) | <0.001 | 4/67 (6.0) | 2/1032 (0.2) | <0.001 |
| Disseminated intravascular coagulation | 1/1099 (0.1) | 0/926 (0.0) | 1/173 (0.6) | 0.157 | 1/67 (1.5) | 0/1032 (0.0) | 0.061 |
| Rhabdomyolysis | 1/1099 (0.1) | 1/926 (0.1) | 0/173 (0.0) | 1.000 | 0/67 (0.0) | 1/1032 (0.1) | 1.000 |
| Pneumonia | 869/1099 (79.1) | 705/926 (76.1) | 164/173 (94.8) | <0.001 | 59/67 (88.1) | 810/1032 (78.5) | 0.087 |
| Time from the initial diagnosis to developing pneumonia (days) |  |  |  |  |  |  |  |
| Median, interquartile range | 0.0 (0.0–2.0) | 0.0 (0.0–2.0) | 1.0 (0.0–3.0) | <0.001 | 1.0 (0.0–5.0) | 0.0 (0.0–2.0) | 0.001 |
| Range | 0.0 (0.0–60.0) | 0.0 (0.0–47.0) | 1.0 (0.0–60.0) | <0.001 | 1.0 (0.0–60.0) | 0.0 (0.0–47.0) | 0.001 |
| Time from symptom onset to developing pneumonia (days) |  |  |  |  |  |  |  |
| Median, interquartile range | 4.0 (2.0–7.0) | 4.0 (2.0–6.0) | 5.0 (3.0–8.0) | <0.001 | 5.5 (3.0–10.0) | 4.0 (2.0–7.0) | 0.015 |
| Range | 4.0 (0.0–46.0) | 4.0 (0.0–43.0) | 5.0 (0.0–46.0) | <0.001 | 5.5 (0.0–46.0) | 4.0 (0.0–43.0) | 0.015 |
| Supportive treatment – No., % |  |  |  |  |  |  |  |
| Administration of intravenous antibiotics – No., % | 632/1099 (57.5) | 493/926 (53.2) | 139/173 (80.3) | <0.001 | 60/67 (89.6) | 572/1032 (55.4) | <0.001 |
| Administration of oseltamivir – No., % | 393/1099 (35.8) | 313/926 (33.8) | 80/173 (46.2) | 0.002 | 36/67 (53.7) | 357/1032 (34.6) | 0.002 |
| Administration of antifungal medications – No., % | 30/1099 (2.7) | 17/926 (1.8) | 13/173 (7.5) | <0.001 | 8/67 (11.9) | 22/1032 (2.1) | <0.001 |
| Administration of systemic corticosteroids – No., % | 204/1099 (18.6) | 127/926 (13.7) | 77/173 (44.5) | <0.001 | 35/67 (52.2) | 169/1032 (16.4) | <0.001 |
| Maximal daily dose of corticosteroids (mg/kg) | 1.5 (0.7–40.0) | 1.0 (0.6–40.0) | 30.0 (1.0–40.0) | 0.014 | 1.6 (1.0–35.0) | 1.5 (0.6–40.0) | 0.505 |
| Oxygen therapy – No., % | 418/1099 (38.0) | 304/926 (32.8) | 114/173 (65.9) | <0.001 | 58/67 (86.6) | 360/1032 (34.9) | <0.001 |
| Mechanical ventilation – No., % | 67/1099 (6.1) | 0/926 (0.0) | 67/173 (38.7) | <0.001 | 40/67 (59.7) | 27/1032 (2.6) | <0.001 |
| Invasive | 24/1099 (2.2) | 0/926 (0.0) | 24/173 (13.9) | <0.001 | 24/67 (35.8) | 0/1032 (0.0) | <0.001 |
| Non-invasive | 56/1099 (5.1) | 0/926 (0.0) | 56/173 (32.4) | <0.001 | 29/67 (43.3) | 27/1032 (2.6) | <0.001 |
| **Use of extracorporeal membrane oxygenation – No., %** | 5/1099 (0.5) | 0/926 (0.0) | 5/173 (2.9) | <0.001 | 5/67 (7.5) | 0/1032 (0.0) | <0.001 |
| **Use of continuous renal replacement therapy – No., %** | 9/1099 (0.8) | 0/926 (0.0) | 9/173 (5.2) | <0.001 | 8/67 (11.9) | 1/1032 (0.1) | <0.001 |
| **Use of intravenous immunoglobulin – No., %** | 143/1099 (13.0) | 86/926 (9.3) | 57/173 (32.9) | <0.001 | 27/67 (40.3) | 116/1032 (11.2) | <0.001 |
| **Intensive care unit admission – No., %** | 55/1099 (5.0) | 22/926 (2.4) | 33/173 (19.1) | <0.001 | 55/67 (82.1) | 0/1032 (0.0) | <0.001 |
| **Clinical outcomes** |  |  |  |  |  |  |  |
| Discharge from hospital | 55/1099(5.0) | 50/926(5.4) | 5/173 (2.9) | 0.230 | 1/67(1.5) | 54/1032 (5.2) | 0.249 |
| Death | 15/1099 (1.4) | 1/926 (0.1) | 14/173 (8.1) | <0.001 | 15/67 (22.4) | 0/1032 (0.0) | <0.001 |
| Recovered | 9/1099 (0.8) | 7/926 (0.8) | 2/173 (1.2) | 0.639 | 0/67 (0.0) | 9/1032 (0.9) | 1.000 |
| Staying in hospital | 1029/1099 (93.6) | 875/926 (94.5) | 154/173 (89.0) | 0.011 | 51/67 (76.1) | 978/1032 (94.8) | <0.001 |

### Clinical outcomes

The percentages of patients being admitted to the ICU, requiring invasive ventilation and death were 5.00%, 2.18% and 1.36%, respectively. This corresponded to 67 (6.10%) of patients having reached to the composite endpoint (**Table 3**).

Results of the univariate competing risk model are shown in **Table E1** in *Supplementary Appendix*. Severe pneumonia cases (SDHR, 9.803; 95%CI, 4.06 to 23.67), leukocyte count greater than 4,000/mm^3^ (SDHR, 4.01; 95%CI, 1.53 to 10.55) and interstitial abnormality on chest X-ray (SDHR, 4.31; 95%CI, 1.73 to 10.75) were associated with the composite endpoint (**Fig. 2**, see **Table E2** in *Supplementary Appendix*). Sensitivity analyses are shown in **Figure E2** in *Supplementary Appendix*.

**Figure 2.**
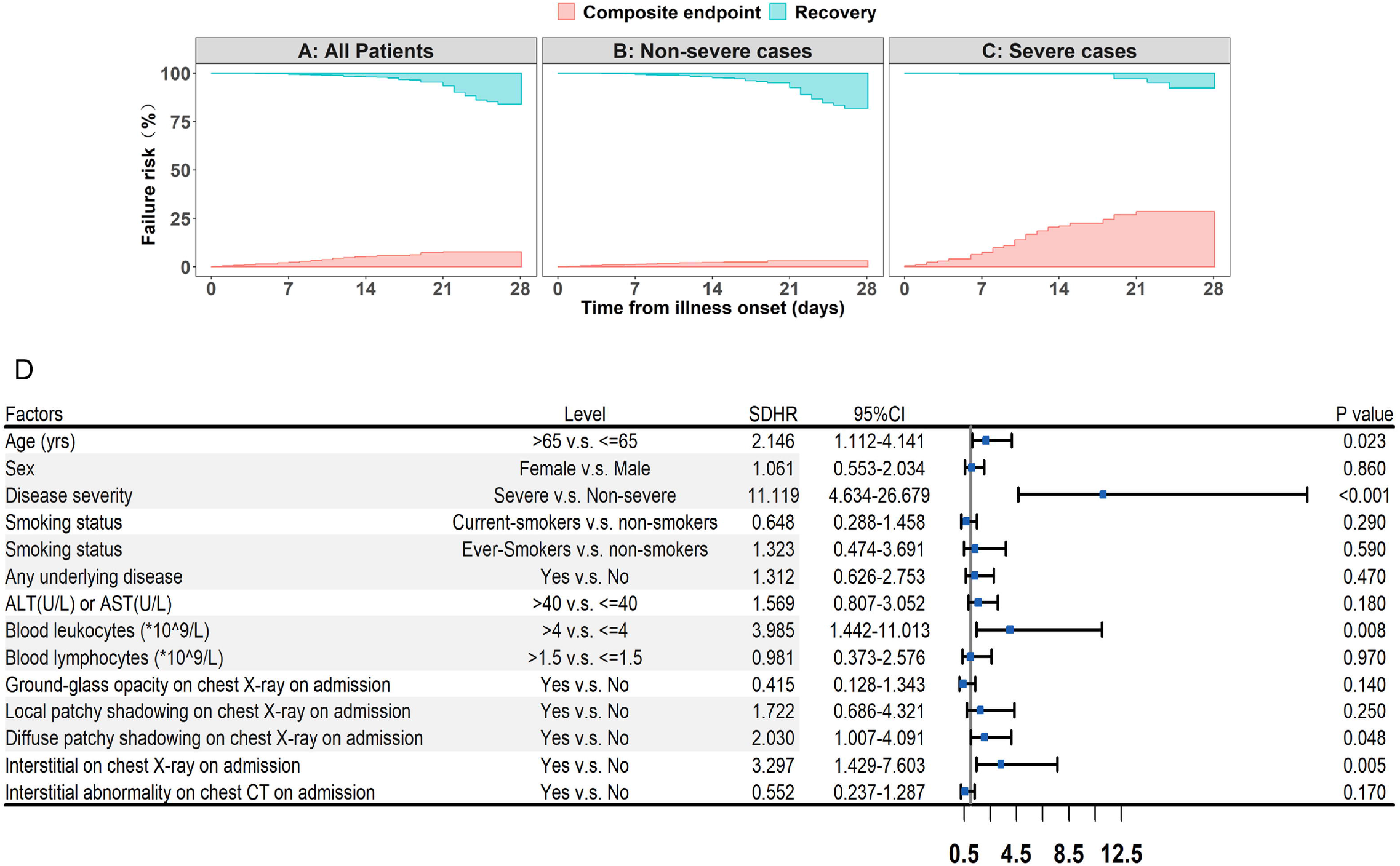
The risk and the percentage of patients with 2019-nCoV ARD who reached to the composite endpoint. Figure 2-A. The risk of reaching to the composite endpoint for all patients with 2019-nCoV ARD Figure 2-B. The risk of reaching to the composite endpoint for non-severe cases Figure 2-C. The risk of reaching to the composite endpoint for severe cases Figure 2-D. Shown are the stratification by age, Sex, disease severity, smoking status, underlying disease, alanine or aspartate aminotransferase levels, blood leukocyte count, blood lymphocyte count, blood platelet count, ground-glass opacity on chest X-ray on admission, local patchy shadowing on chest X-ray on admission, diffuse patchy shadowing on chest X-ray on admission, interstitial abnormality on chest X-ray on admission, interstitial abnormality on chest computed tomography on admission. 2019-nCoV ARD: 2019 novel coronavirus acute respiratory disease

## Discussion

This study has shown that fever occurred in only 43.8% of patients with 2019-nCoV ARD on presentation but developed in 87.9% following hospitalization. Severe pneumonia occurred in 15.7% of cases. No radiologic abnormality was noted on initial presentation in 23.9% and 5.2% of severe and non-severe cases respectively while diarrhea was uncommon. The median incubation period of 2019-nCoV ARD was 3.0 days and it had a relatively lower fatality rate than SARS-CoV and MERS-CoV. Disease severity independently predicted the composite endpoint.

Our study provided further evidence of human-to-human transmission. Around only 1% of patients had a direct contact with wildlife, while more than three quarters were local residents of Wuhan, or had contacted with people from Wuhan. Most cases were recruited after January 1^st^, 2020.

These findings echoed the latest reports, including the outbreak of a family cluster [4], transmission from an asymptomatic individual [6] and the three-phase outbreak patterns [8]. Our study cannot preclude the presence of ‘super-spreaders’. The median incubation period was shorter than a recent report of 425 patients (3.0 days vs. 5.2 days) [8]. Our findings have provided evidence from a much larger sample size to guide the duration of quarantine for close contacts.

Importantly, the routes of transmission might have contributed considerably to the rapid spread of 2019-nCoV. Conventional routes of transmission of SARS-CoV, MERS-CoV and highly pathogenic influenza consisted of the respiratory droplets and direct contact [17-19]. According to our latest pilot experiment, 4 out of 62 stool specimens (6.5%) tested positive to 2019-nCoV, and another four patients in a separate cohort who tested positive to rectal swabs had the 2019-nCoV being detected in the gastrointestinal tract, saliva or urine (see **Tables E3-E4** in *Supplementary Appendix*). In a case with severe peptic ulcer after symptom onset, 2019-nCoV was directly detected in the esophageal erosion and bleeding site (Hong Shan and Jin-cun Zhao, personal communication). Collectively, fomite transmission might have played a role in the rapid transmission of 2019-nCoV, and hence hygiene protection should take into account the transmission via gastrointestinal secretions. These findings will, by integrating systemic protection measures, curb the rapid spread worldwide.

We have adopted the term 2019-nCoV ARD which has incorporated the laboratory-confirmed symptomatic cases without apparent radiologic manifestations. Pneumonia was not mandatory for inclusion. 20.9% patients have isolated 2019-nCoV infection before or without the development of viral pneumonia. Our findings advocate shifting the focus to identifying and managing patients at an earlier stage, before disease progression.

In concert of recent publications [1,8,12], the clinical characteristics of 2019-nCoV ARD mimicked those of SARS-CoV. Fever and cough were the dominant symptoms whereas gastrointestinal symptoms were rare, suggesting the difference in viral tropism as compared with SARS-CoV, MERS-CoV and influenza [20-22]. Notably, fever occurred in only 43.8% of patients on initial presentation and developed in 87.9% following hospitalization. Absence of fever in 2019-nCoV ARD is more frequent than in SARS-CoV (1%) and MERS-CoV infection (2%) [19] and such patients may be missed if the surveillance case definition focused heavily on fever detection [14]. Consistent with two recent reports [1,12], lymphopenia was common and, in some cases, severe. However, based on a larger sample size and cases recruited throughout China, we found a markedly lower case fatality rate (1.4%) as compared with that reportedly recently [1,12]. The fatality rate was lower (0.88%) when incorporating additional pilot data from Guangdong province (N=603) where effective prevention has been undertaken (unpublished data). Our findings were consistent with the national official statistics, reporting the mortality of 2.01% in China out of 28,018 cases as of February 6^th^, 2020 [11,23]. Early isolation, early diagnosis and early management might have collectively contributed to the marked reduction in mortality in Guangdong. Furthermore, dilution of health workforce as a result of central management (i.e., Wuhan JinYinTan Hospital) might have led to increased mortality rate. These findings will inform the mass public, clinicians and policy makers the true transmissability of 2019-nCoV which has resulted in a major social panic.

Our study has stratified patients with 2019-nCoV ARD based on the severity on admission according to international guidelines [15]. Severe cases had significantly higher risk of reaching the composite endpoint. The risk factors indicated the importance of taking into account the disease severity, laboratory findings, chest imaging findings in practice. The applicability of MuLBSTA score, an early warning model for predicting mortality in viral pneumonia, warrants further validation [25].

Despite the markedly high phylogenetic homogeneity as compared with SARS-CoV, there are some clinical characteristics that differentiated 2019-nCoV from SARS-CoV, MERS-CoV, and seasonal influenza which have been more common in respiratory out-patient clinics and wards. **Table E5** in *Supplementary Appendix* highlights the defining characteristics of these viruses, enabling clinicians to differentiate these diagnoses.

Our study has some notable limitations. First, some cases had incomplete documentation of the exposure history, symptoms and laboratory testing given the variation in the structure of electronic database among different participating site and the urgent timeline for data extraction. Some cases were diagnosed in out-patient settings where medical information was briefly documented and incomplete laboratory testing was applied. There was a shortage of infrastructure and training of medical staff in non-specialty hospitals, which has been aggravated by the burn-out of local medical staff in milieu of a surge of cases. Second, because many patients still remained in the hospital, we did not compare the 28-day rate of the composite endpoint. To mitigate the potential bias, we have applied the competing-risk model for analysis. Third, we might have missed asymptomatic or mild cases managed at home, and hence our cohort might represent the more severe end of 2019-nCoV ARD. However, there were a minority of patients who had no apparent radiologic manifestations, suggesting that we had included patients at the early stage of disease. Last, we took reference on the existing international guideline to define the severity of 2019-nCoV because of its global recognition [15].

In summary, 2019-nCoV elicits a rapid spread of outbreak with human-to-human transmission, with a median incubation period of 3 days and a relatively low fatality rate. Absence of fever and radiologic abnormality occurs in a substantial proportion of patients on initial presentation while diarrhea is uncommon. The disease severity is an independent predictor of poor outcome. Stringent and timely epidemiological measures are crucial to curb the rapid spread. Ongoing efforts are needed to explore for an effective therapy (i.e., protease inhibitors, remdesivir, β interferon) for this emerging acute respiratory infection.

## Data Availability

Data will be made available upon request to the corresponding author.

## Author’s contribution

W. J. G., J. X. H., W. H. L., C. Q. O., P. Y. C., L. J. L., G. Z., K. Y. Y., B. D., and N. S. Z. participated in study design; C. Q. O., P. Y. C., W. J. G., and W. H. L. performed data analysis; Z. Y. N., L. L., H. S., C. L. L., L. J. L., G. Z., K. Y. Y., B. D., R. C. C., C. L. T., T. W., J. X., S. Y. L., J. L. W., Z. J. L., Y. H., Y. X. P., L. W., Y. L., Y. H. H., P. P., J. M. W., J. Y. L., Z. C., G. L., Z.J. Z., S. Q. Q., J. L., C. J. Y., S. Y. Z., and N. S. Z. recruited patients; W. J. G., J. X. H., W. H. L., D. S. C. H., and N. S. Z. drafted the manuscript; W. J. G., J. X. H., W. H. L., C. Q. O., Z. Y. N., L. L., H. S.,C. L. L., D. S. C. H., L. J. L., G. Z., K. Y. Y., B. D., and N. S. Z. were responsible for study conception; all authors provided critical review of the manuscript and approved the final draft for publication.

## Acknowledgment

We thank the hospital staff (see Supplementary Appendix for a full list of the staff) for their efforts in recruiting patients. We are indebted to the coordination of Drs. Zong-jiu Zhang, Ya-hui Jiao, Bin Du, Xin-qiang Gao and Tao Wei (National Health Commission), Yu-fei Duan and Zhi-ling Zhao (Health Commission of Guangdong Province), Yi-min Li, Zi-jing Liang, Nuo-fu Zhang, Shi-yue Li, Qing-hui Huang, Wen-xi Huang and Ming Li (Guangzhou Institute of Respiratory Health) which greatly facilitate the collection of patient’s data. Special thanks are given to the statistical team members Prof. Zheng Chen, Drs. Dong Han, Li Li, Zheng Chen, Zhi-ying Zhan, Jin-jian Chen, Li-jun Xu, Xiao-han Xu (State Key Laboratory of Organ Failure Research, Department of Biostatistics, Guangdong Provincial Key Laboratory of Tropical Disease Research, School of Public Health, Southern Medical University). We also thank Li-qiang Wang, Wei-peng Cai, Zi-sheng Chen, Chang-xing Ou, Xiao-min Peng, Si-ni Cui, Yuan Wang, Mou Zeng, Xin Hao, Qi-hua He, Jing-pei Li, Xu-kai Li, Wei Wang, Li-min Ou, Ya-lei Zhang, Jing-wei Liu, Xin-guo Xiong, Wei-juna Shi, San-mei Yu, Run-dong Qin, Si-yang Yao, Bo-meng Zhang, Xiao-hong Xie, Zhan-hong Xie, Wan-di Wang, Xiao-xian Zhang, Hui-yin Xu, Zi-qing Zhou, Ying Jiang, Ni Liu, Jing-jing Yuan, Zheng Zhu, Jie-xia Zhang, Hong-hao Li, Wei-hua Huang, Lu-lin Wang, Jie-ying Li, Li-fen Gao, Jia-bo Gao, Cai-chen Li, Xue-wei Chen, Jia-bo Gao, Ming-shan Xue, Shou-xie Huang, Jia-man Tang, Wei-li Gu, Jin-lin Wang (Guangzhou Institute of Respiratory Health) for their dedication to data entry and verification.

## Reference

1. Huang C, Wang Y, Li X, et al. Clinical features of patients with 2019 novel coronavirus in Wuhan, China. Lancet. 2020; doi: 10.1016/S0140-6736(20)30183-5

2. Lu R, Zhao X, Li J, et al. Genomic characterization and epidemiology of 2019 novel coronavirus: implications of virus origins and receptor binding. Lancet. 2020; doi: 10.1016/S0140-6736(20)30251-8

3. Zhu N, Zhang D, Wang W, et al. A novel coronavirus from patients with pneumonia in China, 2019. N Engl J Med. 2020; doi: 10.1056/NEJMoa2001017

4. Chan JF, Yuan S, Kok KH, et al. A familial cluster of pneumonia associated with the 2019 novel coronavirus indicating person-to-person transmission: a study of a family cluster. Lancet. 2020; doi: 10.1016/S0140-6736(20)30154-9

5. Phan LT, Nguyen TV, Luong QC, et al. Importation and human-to-human transmission of a novel coronavirus in Vietnam. N Engl J Med. 2020; doi: 10.1056/NEJMc2001272

6. Rothe C, Schunk M, Sothmann P, et al. Transmission of 2019-nCoV infection from an asymptomatic contact in Germany. N Engl J Med. 2020; doi: 10.1056/NEJMc2001468

7. Wu JT, Leung K, Leung GM. Nowcasting and forecasting the potential domestic and international spread of the 2019-nCoV outbreak originating in Wuhan, China: A modeling study. Lancet. 2020; doi: 10.1016/S0140-6736(20)30260-9

8. Li Q, Guan X, Wu P, et al. Early transmission dynamics in Wuhan, China, of novel coronavirus-infected pneumonia. N Engl J Med. 2020; doi: 10.1056/NEJMoa2001316

9. WHO main website. https://www.who.int (accessed February 5th, 2020)

10. Holshue ML, DeBolt C, Lindquist S, et al. First case of 2019 novel coronavirus in the United States. N Engl J Med. 2020; doi: 10.1056/NEJMoa2001191

11. National Health Commission of the People’s Republic of China. http://www.nhc.gov.cn (Assessed on February 6th, 2020)

12. Chen N, Zhou M, Dong X, et al. Epidemiological and clinical characteristics of 99 cases of 2019 novel coronavirus pneumonia in Wuhan, China: a descriptive study. Lancet. 2020. doi: 10.1016/S0140-6736(20)30211-7

13. New coronavirus pneumonia prevention and control program (2nd ed.) (in Chinese). 2020 (http://www.nhc.gov.cn/jkj/s3577/202001/c67cfe29ecf1470e8c7fc47d3b751e88.shtml). (accessed February 6th, 2020)

14. WHO. Clinical management of severe acute respiratory infection when Novel coronavirus (nCoV) infection is suspected: interim guidance. Jan 28, 2020. https://www.who.int/internal-publications-detail/clinical-management-of-severe-acute-respiratory-infection-when-novel-coronavirus-(ncov)-infection-is-suspected (accessed February 5th, 2020)

15. Metlay JP, Waterer GW, Long AC, et al. Diagnosis and treatment of adults with community-acquired pneumonia: An official clinical practice guideline of the American Thoracic Society and Infectious Disease Society of America. Am J Respir Crit Care Med. 2019; 200:e45-e67

16. Laboratory diagnostics for novel coronavirus. WHO 2020 (https://www.who.int/health-topics/coronavirus/laboratory-diagnostics-for-novel-coronavirus) (accessed February 6th, 2020)

17. Lei H, Li Y, Xiao S, et al. Routes of transmission of influenza A H1N1, SARS CoV, and norovirus in air cabin: Comparative analyses. Indoor Air. 2018;28:394–403

18. Otter JA, Donskey C, Yezli S, et al. Transmission of SARS and MERS coronaviruses and influenza virus in healthcare settings: the possible role of dry surface contamination. J Hosp Infect. 2016;92:235–50

19. Zumla A, Hui DS, Perlman S. Middle East respiratory syndrome. Lancet. 2015;386:995–1007

20. Leung WK, To KF, Chan PK, et al. Enteric involvement of severe acute respiratory syndrome-associated coronavirus infection. Gastroenterology. 2003;125:1011–7

21. Assiri A, McGeer A, Perl TM, et al. Hospital outbreak of Middle East respiratory syndrome coronavirus. N Engl J Med. 2013;369:407–16

22. Minodier L, Charrel RN, Ceccaldi PE, et al. Prevalence of gastrointestinal symptoms in patients with influenza, clinical significance, and pathophysiology of human influenza viruses in faecal samples: what do we know? Virol J. 2015;12:215

23. World Health Organization. Novel Coronavirus (2019-nCoV) situation reports. https://www.who.int/emergencies/diseases/novel-coronavirus-2019/situation-reports/ (Assessed on February 6th, 2020)

24. Guo L, Wei D, Zhang X, et al. Clinical features predicting mortality risk in patients with viral pneumonia: the MuLBSTA score. Front Microbiol 2019; 10: 2752

